# “*She helped from the first minute to the last*” – experiences of respectful maternal and newborn care during the COVID-19 pandemic in Nampula Province, Mozambique

**DOI:** 10.1101/2024.03.19.24304557

**Authors:** Megan M Lydon, Joaquim Vilanculos, Carter Crew, Américo Barata, Emily Keyes

## Abstract

Pandemic-related health service adaptations raised concerns about provision of quality, respectful maternity care globally. Despite this, little research has focused on the experiences of those using intrapartum care during this time. This study aimed to elevate the voices and document the experiences of birthing people in Nampula Province, Mozambique during the COVID-19 pandemic. We conducted a longitudinal qualitative study from March-August 2021 and present an analysis of the 17 follow-up in-depth interviews conducted with participants who had a vaginal live birth. Interviews explored participants’ experience of labor and delivery care. They were conducted in Makua and Portuguese, audio-recorded, transcribed and translated. We applied thematic content analysis. Overall, participants did not express major concerns about COVID-19 or related service adaptations when describing their experiences of intrapartum care. Some noted its negative effects on elements of respectful care such as restricting birth companions. Overcrowding became more concerning due to the threat of infection. While unclear if affected by the pandemic, all participants who gave birth at a health facility reported experiencing at least one form of mistreatment, some recounting threats of cesarean delivery. Most explained that they and their newborns received care without their consent, especially regarding enemas and episiotomies. At the same time, respondents described a range of intrapartum experiences that included both respectful and disrespectful care. Most recalled positive verbal communication with their providers and many described receiving continuous attentive care. Participants explained that their satisfaction with childbirth services was tied to their birth outcome and their experience of respectful care. The findings indicate that steadfast commitments to quality care are critical to ensure families benefit from high-quality, respectful care at all times. The ramifications of the COVID-19 pandemic were limited but nonetheless signal a need for tighter connections between maternal health and emergency preparedness stakeholders.

## Introduction

In response to the COVID-19 pandemic, health systems around the world quickly adapted service delivery to prevent viral transmission and manage higher caseloads. Some of these adaptations raised concerns about the ability to offer quality, respectful maternity services.(1) Early pandemic reports documented disrespect and poor quality care including performing unnecessary cesarean deliveries and other obstetric interventions to avoid transmission from laboring clients with COVID-19, abandoning laboring clients, disallowing birth companions, and separating postpartum clients from their newborns.(2, 3)

The World Health Organization defines quality of care as encompassing both the *provision* and the *experience* of care.(4) Experience of care has been associated with childbirth satisfaction and selection of delivery location.(5) Further, respectful, dignified care is a human right.(1) During shocks to a health system, respectful care may be sacrificed while focusing on managing the shock, such as the COVID-19 pandemic. As such, it is critical to document experience of care during health system shocks to hold systems accountable, to offer learnings for future health shocks, and to inform routine service delivery such that care during these periods of stress may highlight underlying service delivery inadequacies.

Limited research has continued to explore the effects of the COVID-19 pandemic on client experience of childbirth care. These studies, largely set in high-income countries, affirm early reports. Restrictions on partner access during childbirth were observed in Ireland, Italy, and Spain.(6, 7) Clients perceived receiving lower quality of care in Spain and reported lower childbirth satisfaction in the United States of America during the pandemic compared to clients pre-pandemic.(8, 9) There was an increased use of obstetric interventions reported in Canada and Italy.(10, 11) A 12-country European study identified gaps in informed consent, breastfeeding support and infection prevention practices.(12) A global survey, comprising mainly high- and middle-income respondents, found that 17% of maternal and newborn health workers reported that their ability to provide respectful care during the COVID-19 pandemic was somewhat or substantially lower than before the pandemic.(13)

Less is known about the pandemic experiences of childbirth in low-income countries. A study conducted in northwest Ethiopia during COVID-19 found that 66% of providers did not explain procedures to birthing clients prior to performing them and 64% did not obtain consent for procedures during labor and delivery but it is unclear how these rates compare to those pre-pandemic.(14) This research offers important insights about provider adherence to respectful maternity care standards, however, there remains a notable gap in documenting client experience throughout the pandemic. Following this, the current study aimed to explore client experience of intrapartum care during the pandemic in Mozambique.

## Study setting

There have been steady increases in institutional delivery rates in Mozambique from 50% to 64% between 2003(15) and 2020(16). At the same time, maternal and neonatal mortality and morbidity remain high. The maternal mortality ratio was estimated at 408 deaths/100,000 live births(17) and the neonatal mortality rate at 24 deaths/1,000 live births in the last demographic and health surveys.(18) The national emergency obstetric care assessment of 2012 revealed gaps in service delivery such that 54% of health centers had not provided parenteral anticonvulsants in the last three months, 46% had not removed retained products of conception and 43% had not administered parenteral antibiotics.(19) Cause-specific fatality rates were high with a 7.0% mortality rate among those with postpartum sepsis, 5.2% fatality rate among those with a ruptured uterus and a 3.2% rate among those who experienced postpartum hemorrhage.(19) As such, critical efforts to improve quality of care have emerged including Mozambique’s establishment of a Department of Management and Quality Assurance, as well as the development and implementation of a quality improvement supervision guide for health facilities.(20)

The first case of COVID-19 in Mozambique was confirmed on March 22, 2020, and quickly prompted large-scale lockdowns. The government of Mozambique announced a State of Emergency on March 31, 2020, with accompanying policies to limit COVID-19 spread. While confirmed cases remained relatively low in Mozambique, health system adaptations were instituted to prevent interpersonal transmission.(21)

This study took place in Nampula Province, located in northern Mozambique. The province is the most populous in the country, home to over 6 million inhabitants, constituting 20% of the national population.(22) Nampula has a 6.1 total fertility rate and the highest adolescent birth rate in the country (17). Nampula lags behind the national institutional delivery rate whereby 52% of women delivered at a health facility in 2020.(18) Research has shown that there was a drop in facility deliveries at the onset of the COVID-19 pandemic in Nampula Province but that rates returned to pre-pandemic levels by April 2021.(23) This study focuses on Erati and Nacala-Porto districts, representing a rural and urban area, respectively. Both are home to the Makhua-speaking population but differ in religious representation. In Erati, 53% identify as Catholic and 40% as Muslim whereas those in Nacala-Porto predominantly identify as Muslim.(24, 25) As a coastal region, Nacala-Porto sees a convergence of different populations and working classes.

## Methods

### Design and data collection

This was a longitudinal qualitative study whereby in-depth interviews (IDIs) were conducted with participants while pregnant and again postpartum. Participants were recruited with support from health providers and community health workers in the two study districts. Those eligible for the study were 18 years or older, currently pregnant, had attended at least one antenatal care (ANC) visit for their current pregnancy, and resided in a study district. We aimed to include an equal number of participants from each study district with a variety of delivery location intentions (at the health facility, outside of the health facility, uncertain), as the primary goal of the study was to investigate care-seeking behavior. To have sufficient data to reach 80% saturation, we aimed to include approximately eight participants in each of the three profiles.(26) The secondary objective of the research was to explore the experience of care during the pandemic, which is the focus of this paper.

We conducted 24 antenatal IDIs March 2-5, 2021, and followed-up with participants after their expected delivery dates to conduct postpartum IDIs. Those who had a non-surgical live birth were eligible for follow-up to focus on the experience of routine intrapartum care. Among the 24 initial participants, 20 were eligible for the postpartum interview; of these, two were lost-to-follow-up and one discontinued participation. We conducted 17 postpartum IDIs August 17-27, 2021, using a semi-structured interview guide exploring participants’ experience of intrapartum care (Table 1). Interviews were conducted by two female research assistants from Nampula who were given training in research ethics and qualitative research. The research assistants were current medical students, allowing them to understand intrapartum processes, and spoke the local language. Interviews took place at a location chosen by the participant, often their home or local health facility, and were conducted in Makua or Portuguese, depending on participant preference. Postpartum interviews lasted 75 minutes on average. IDIs were audio-recorded, directly transcribed into Portuguese, and translated into English.

**Table 1.**

| Post-partum IDI domains |
| --- |
| Initiation of labor and delivery |
| Infection concerns/COVID-19 |
| Interpersonal support and interactions |
| Procedures/clinical care |
| Comfort measures & respectful care |
| Perceptions of intrapartum care |

### Data analysis

We applied thematic content analysis to identify themes and sub-themes. A codebook was developed *a priori* based on the domains of the IDI guide (Table 1). It was further refined through immersive reading of initial transcripts. Coding was conducted by one analyst using Nvivo v.12.(27) Data reduction and synthesis took place through the creation of results matrices. Analytical memos were developed and summarized by two female American analysts. Results were shared with the Mozambique research team for validation. The results presented here center on participants’ experience of intrapartum care.

### Ethical considerations

This study was approved by FHI 360’s Office of International Research Ethics and the Nampula Provincial Delegation of the National Institute of Health in Mozambique. Written informed consent was obtained from all participants.

## Results

### Participant characteristics

Among the 17 postpartum participants, 8 were located in Erati and 9 in Nacala-Porto. They ranged in age from 18-32 years, with an average age of 24. Nearly all were married. Participants in Erati tended to have primary school education and most identified as farmers. Most of those in Nacala-Porto had attended at least some secondary school, with some reporting engaging in formal employment (Table 2). Participants reported birthing between 0 and 5 times prior to the study, such that 35% experienced their first birth within the context of this study. A couple of participants had previously experienced miscarriage and a couple had children who had died. All births took place between March and August 2021. All participants identified as women and described their partners as men, as such, the associated pronouns are used throughout the findings.

**Table 2.**
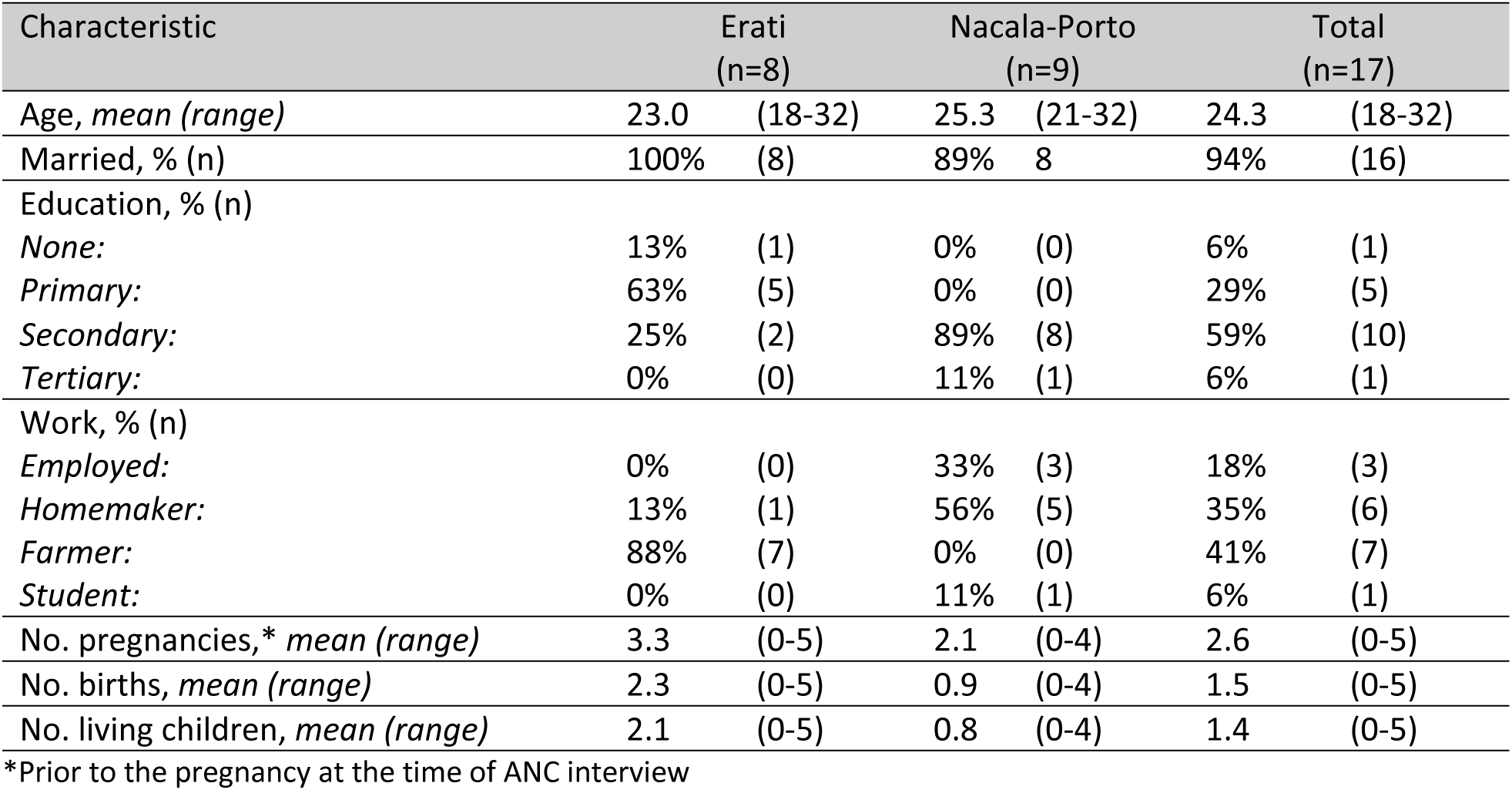
Sociodemographic characteristics of participants, by district.

### Experience of intrapartum care

Among the 17 postpartum participants, one delivered at home, two delivered on their way to the health facility and one was referred between two health facilities. All others gave birth at the first health facility they attended, all of which were publicly operated facilities. The experiences of the participant who birthed at home are included only when relevant (e.g., they brought their newborn into the health facility so descriptions of newborn care experience, general perceptions of what constitutes a positive or negative birth experience, and reasons for level of birth satisfaction are included).

### Health facility admissions

Most participants reported being attended to “*immediately*” or “*very fast*” upon arrival at the health facility. Several however described encountering delays at the health facility, explaining that providers were busy with other patients, or the room was being cleaned.

Participants consistently reported having their blood pressure and temperature measured at admissions. About half of participants noted their provider took a urine sample or asked them if they were experiencing vaginal bleeding or headaches. Many recounted having their “*belly*” measured, referring to measuring uterine height, and receiving an initial vaginal exam. All participants reported feeling at ease with these initial screening measures such that one explained, “*That was really comfortable and confident for me because it is a normal procedure*.” Among those who did not receive screening tests, one recounted that she had arrived after regular health facility hours so there were not enough providers to do a comprehensive intake, while another explained the providers were busy when she arrived.

### Labor and childbirth

Participants described a range of experiences that included both respectful and disrespectful care. All participants who gave birth at a health facility reported experiencing at least one form of mistreatment during labor and delivery. The results are organized according to Bohren and colleagues’ (2015) typology of the mistreatment of laboring persons during childbirth.(28) We expanded this framework to include a corresponding equivalent typology of *respectful* care during childbirth to capture the positive experiences reported by respondents. Select quotes are featured in Table 3. Some of the experiences described by participants align with more than one theme but have been presented only once for brevity.

**Table 3.**
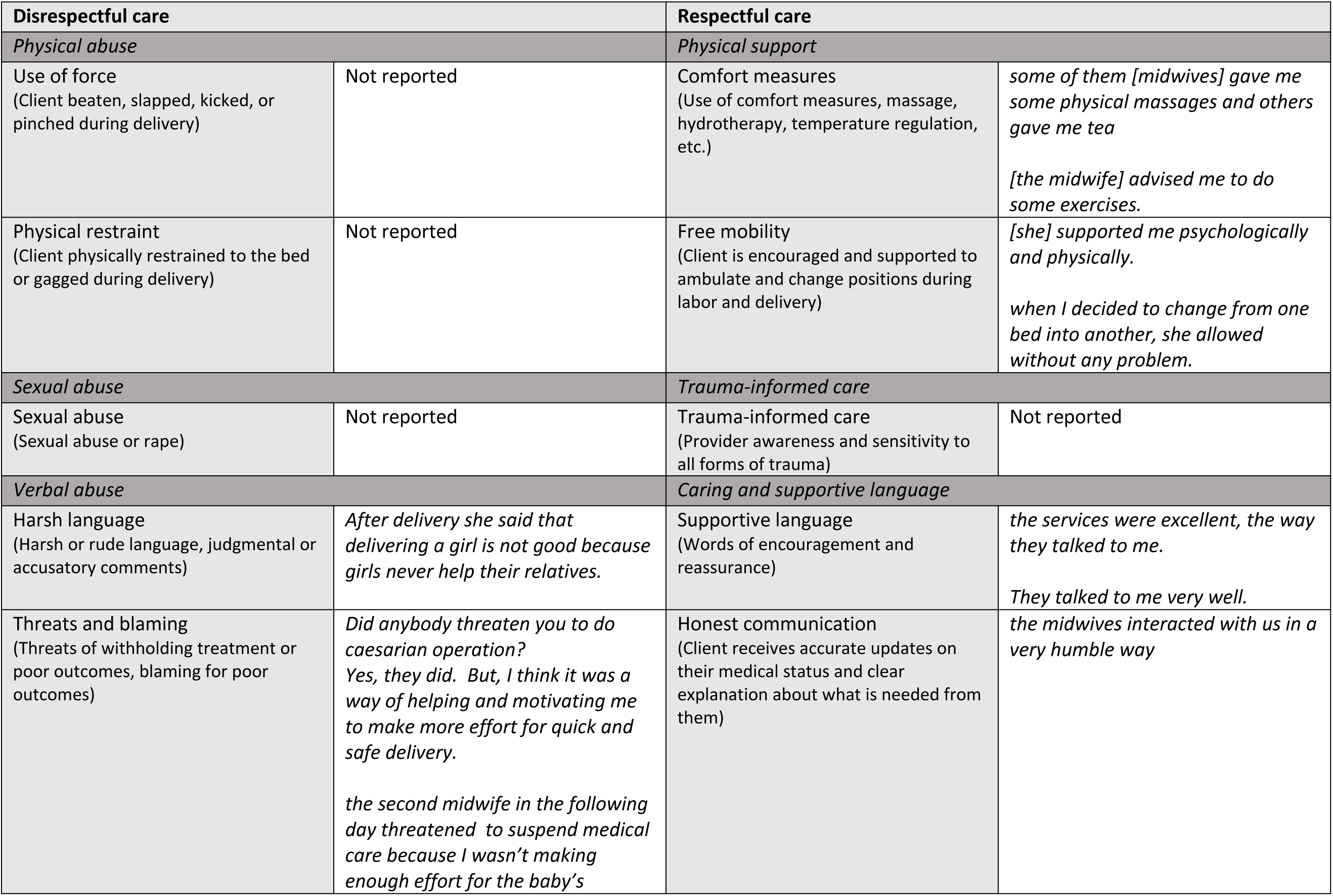
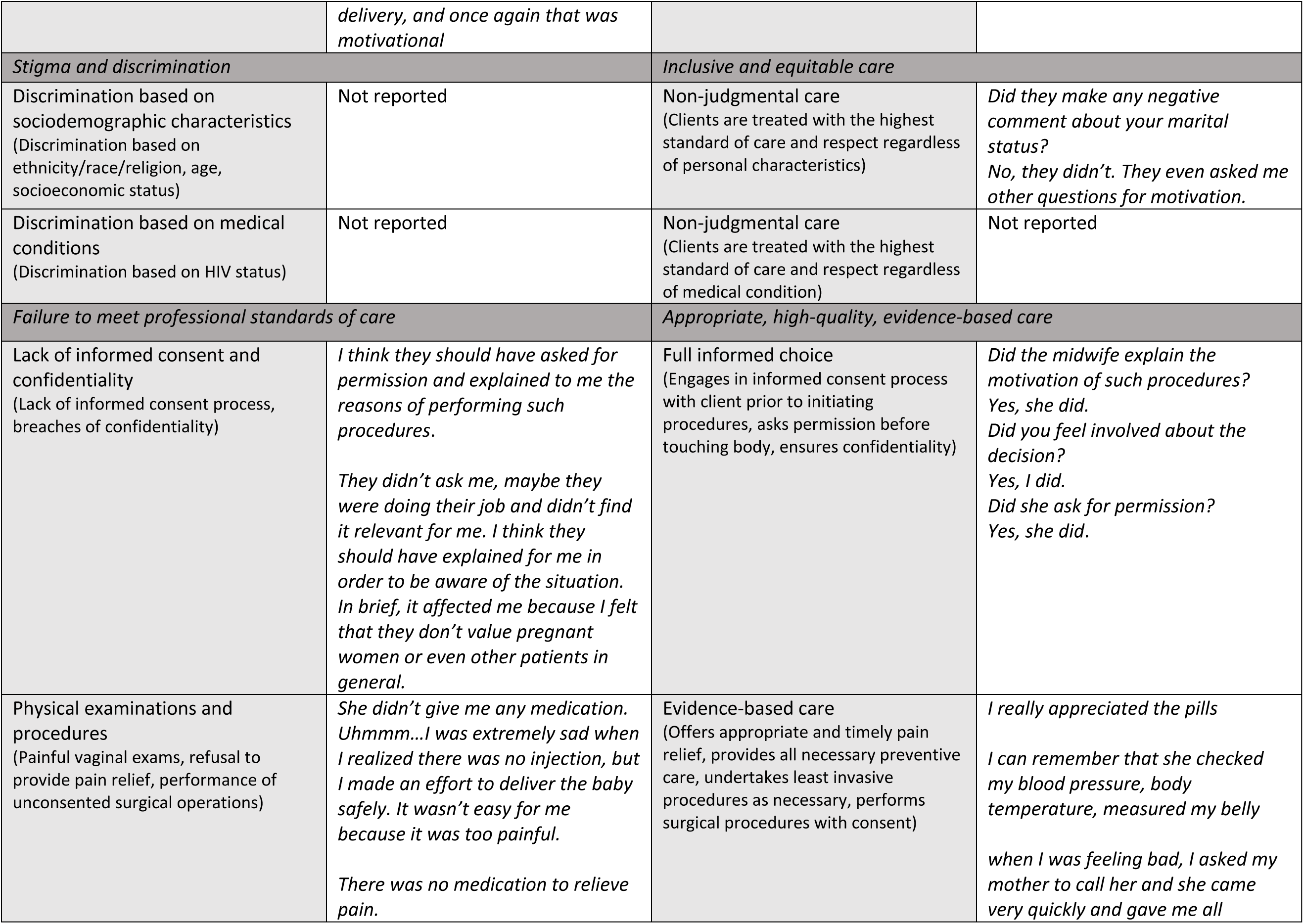
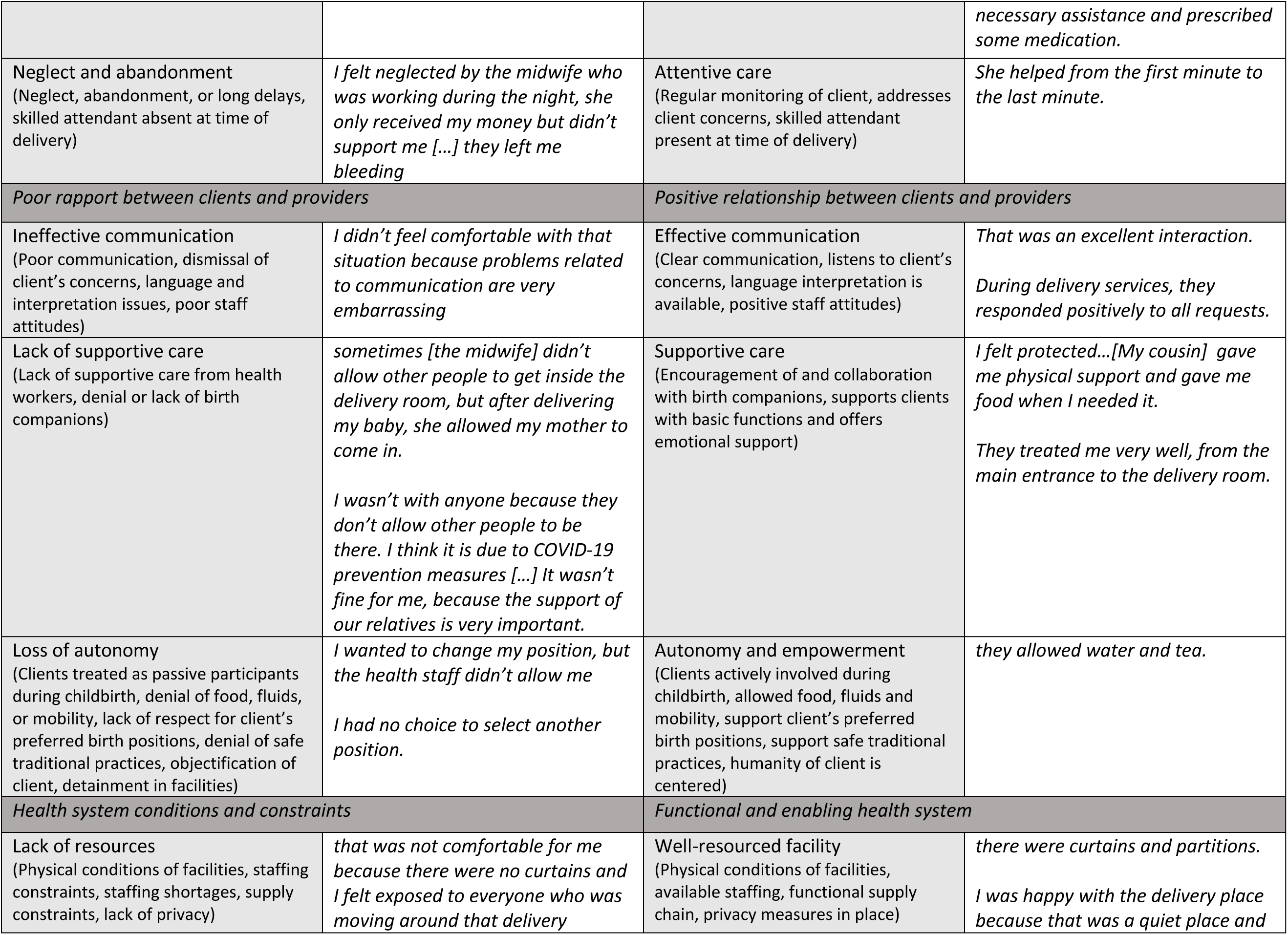
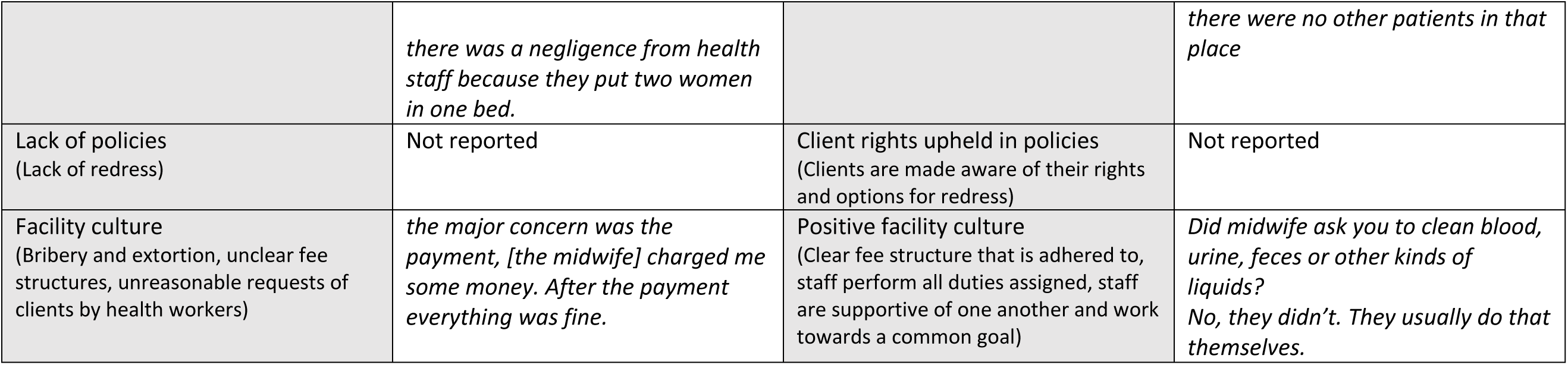
Examples of reported experiences of respect and disrespect during intrapartum care.

#### Physical abuse/ Physical support

There were no reports of physical abuse. In contrast, many participants described receiving physical support from their providers. This included receiving assistance with walking and changing positions in bed to improve comfort and aid in labor progression.

> *They helped me to walk in order to facilitate the delivery process*

Participants appreciated receiving massages to manage labor pain and assistance with personal hygiene. Several said the midwife “*touched my body very well*” and explained that they felt respected when provided with physical support.

#### Verbal abuse/ Caring and supportive language

##### Harsh versus caring language

Most participants recalled positive verbal communication with their providers, saying “*she talked to me very well,*” however, one respondent noted that her midwife shouted at her during labor and delivery. Some expressed surprise by the positive communication they received at the health facility, such that one participant explained, “*[the midwife] treated me very well, they didn’t shout at me, they didn’t even insult me. In some hospitals the health staff usually slap the patients*.” This suggests that participants expected negative interactions at the health facility but discovered better care than anticipated.

##### Threats and blaming

Some participants reported experiences of threats and blaming. A couple recounted that their midwife made derogatory remarks about their baby’s sex.

> *After delivery she said that delivering a girl is not good because girls never help their relatives.*

A few others shared that their provider threatened them with a cesarean delivery, one of whom additionally recalled the providers threatening to withhold care if she did not make more progress. The participants perceived this behavior as acceptable and useful in their birth experience, whereby one said, “*They simply threatened me because I wasn’t making enough effort for the delivery, and then the health staff decided to use this technique in order to encourage me to make more effort for a safe delivery*.” Despite this justification, she later went on to note that “*my least favorite part of the experience was when they threatened me for caesarian operation,*” indicating that while the approach has been normalized, participants were willing to express discontent.

#### Stigma and discrimination/ Inclusive and equitable care

There were no reports of stigma or discrimination directed towards the birthing person. Participants consistently reported that they were not subject to derogatory remarks about their physical appearance, sexual activity, ethnicity, race, tribe, culture, religion, age, marital status, level of education or literacy, economic circumstances, or HIV status. One unmarried participant described that the providers did not make any negative comments about her marital status and “*even asked [her] other questions for motivation*,” highlighting how her providers continued to respectfully and meaningfully engage with her despite her marital status.

#### Failure to meet professional standards of care / Appropriate, high-quality, evidence-based care

##### Informed consent and physical examinations and procedures

Most participants recalled that their provider listened to their fetal heart beats at some point during labor. In almost all cases, participants reported that their provider explained the purpose of this procedure and asked permission before placing the fetoscope on their stomach. All participants said they felt comfortable with the level of privacy during this procedure and most enjoyed the experience saying, “*I liked it too much*” and “*that was an interesting experience!*” such that they felt connected to their child and reassured about their child’s well-being.

We asked participants about their experience of vaginal exams and any major interventions performed during labor and delivery including both evidence-based and non-evidence-based care. To note, the WHO does not recommend pubic shaving, routine episiotomy, or enema on admission.(29) All but one participant who delivered at a health facility received a vaginal exam, most reported receiving an enema or some form of rectal cleaning, about half received episiotomies, and many received stitches to manage vaginal tearing or episiotomy. Several reported receiving some form of “*injection*” to induce or augment labor, however, participants were unsure of the specifics. There were no reports of vaginal shaving. No participants described having an assisted vaginal delivery, an intervention that is recommended when indicated.(30)

Most respondents reported experiences of unconsented care. While most said their provider asked permission before performing a vaginal exam, about half noted that their provider did not explain the purpose of the exam or its necessity. Most participants said their experience of the vaginal exam “*was fine*” or was an “*interesting experience*.” A couple acknowledged the physical discomfort of the procedure such that one explained, “*nobody likes her genitals to be touched, and it is also very uncomfortable*.” Many expressed discomfort with the lack of privacy during vaginal exams. One participant echoed the sentiments shared by others saying, “*They touched my vagina and that wasn’t comfortable for me because there were five people in the delivery room*.”

Several respondents reported not being asked permission or provided with an explanation for interventions performed, particularly for enemas and episiotomies. Participants perceived the procedure of anal cleaning as “*normal*” even when unconsented, saying they felt it contributed to good “*hygiene*.” One participant described the prevalence of unconsented care as standard practice saying, *“they didn’t explain to me the reason for performing such procedures. Health staff usually don’t explain why they have to perform a certain procedure.”* While lack of informed consent may have been typical, a few participants expressed a desire for a different standard of care. One who experienced manual removal of her placenta and rectal cleaning, explained:

> *They didn’t ask me, maybe they were doing their job and didn’t find it relevant for me. I think they should have explained for me in order to be aware of the situation. In brief, it affected me because I felt that they don’t value pregnant women or even other patients in general.*

Regarding newborn care, most participants recounted holding their baby “*immediately*” or within minutes after delivery. A few participants reported holding their newborn for the first time 1-2 hours after delivery, but they did not know what caused this delay. In contrast, one mother was given her baby immediately after delivery but was unable to hold them given her medical condition, explaining, “*[the midwife] didn’t support me, she simply took my baby, cleaned them and gave them to me while I was still weak and bleeding*.” A minority of participants recalled providers performing any procedures on their newborn including suctioning, blood testing, administration of antibiotic eye ointment or vitamin K injection. Among those whose infants received a medical procedure, all noted that their provider first asked their permission but nearly all also said the provider did not explain the purpose or necessity of the intervention. Overall, participants described meeting their child as “*exciting*” and feeling very “*happy*.”

### Pain relief

Several of those who birthed in the health facility were not offered any form of pain relief. Among those who received pain relief, most described being prescribed “*pills*” and one explained she received an unspecified “*injection*.” One participant recounted receiving stitches without anesthesia and described this as the moment that broke her trust in her providers:

> *I got some stitches because the baby was big and heavy, that’s why they had to open my vagina more to allow the removal the baby from there. It was a very painful procedure because they didn’t use anesthesia for that […] I was confident about their skills when they advised and helped me during the delivery process; however, I didn’t feel confident when they enlarged my vagina in order to remove the baby from there and that was extremely painful due to lack of anesthesia.*

While medical options for pain relief were limited, several described how the nursing staff or their birth companions were able to offer comfort measures such as massage, movement, and alternating birth positions such that respondents described the midwife “*moved my body*” and “*advised me to do some exercises*.” Participants were deeply appreciative of such care.

### Neglect versus attentive care

A couple participants reported being ignored or neglected by providers. One justified her experience saying that “*maybe the midwife was busy*.” Another participant was left unattended while hemorrhaging even though she requested assistance:

> *I felt neglected by the midwife who was working during the night, she only received my money but didn’t support me […] they left me bleeding*

Many participants however recounted experiencing continuous, attentive care, such that some explained, “*[the midwife] helped from the first minute to the last minute*.” Further, nearly all felt that their providers responded to their concerns.

*When I was feeling bad, I asked my mother to call [the nurse] and she came very quickly and gave me all necessary assistance and prescribed some medication […] during delivery services [the nurse] was patient and understood my situation*.

#### Poor rapport between clients and providers / Positive relationship between clients and providers

##### Communication

One participant described being unable to communicate in a common language with her providers, with no option for language interpretation, saying, “*I didn’t feel comfortable with that situation because problems related to communication are very embarrassing*.” Participants recounted feeling respected when providers engaged in active listening, were patient and understanding of their circumstances, and approached them with a “*humble*” attitude.

##### Supportive care

Nearly all participants had the support of one or more birth companion during labor and delivery. Respondents strongly appreciated that support, which was seen as complementing the care received from facility staff. They described their birth companions, usually female relatives and traditional birth attendants, as offering motivation, reassurance, and comfort, and providing physical support in the form of comfort measures, mobility assistance and performing daily activities such as washing clothes. Participants felt their birth companions had very positive interactions with the health staff.

In some cases, while birth companions were permitted in the health facility, they were not allowed in the labor and delivery room. Participants believed this to be attributed to COVID-19 prevention measures and/or to ensure privacy for patients. In all cases it was upsetting for the birthing clients.

> *I wasn’t with anyone because they don’t allow other people to be there. I think it is due to COVID-19 prevention measures […] It wasn’t fine for me because the support of our relatives is very important.*

Another couple of participants noted they were limited to one companion in the maternity unit, so they had to choose between the members of their support team which sometimes included their husband, mother, sister, and/or traditional birth attendant.

> *She only allowed the traditional birth attendant. Due to COVID-19, there is a restriction on entry of people. They allow only one person […] My mother was very sad due to my situation. Because my mother wasn’t allowed to get into the delivery room due to COVID-19 restrictions.*

##### Autonomy

The WHO recommends oral fluid and food intake during labor for low-risk clients, as well as a birth position of their choice including upright positions.(29) In this study, most participants remembered being permitted to drink fluids during labor and recalled drinking “*water*” or “*tea*.” At the same time, most reported not being allowed to eat during labor. Several said that this requirement did not affect them negatively with one explaining that she thought “*it is not recommended to eat any food during the delivery process, otherwise it may cause other complications like defecation*.” When asked about their preferred birthing position, respondents exclusively said, “*the lying down position*.” One participant noted, “*I think the position is the only one…you know?*”, indicating a lack of awareness about other potential labor positions.

Most participants recounted feeling supported to birth in their chosen position, the supine position. At the same time, one respondent said she had no choice to select another position, and another described feeling uncomfortable lying down and “*wanted to change the position but the health staff didn’t allow [her]*.” While birth positions were limited, several participants described receiving assistance with walking and physical movement in support of their bodily autonomy.

#### Health system conditions and constraints / Functional and enabling health systems

##### Health facility environment

Most participants felt comfortable with their labor and delivery setting, noting it was “*clean*” and they felt “*happy*.” Despite this, most also described the privacy measures as unsatisfactory. One participant highlighted this saying, “*It was an embarrassing situation for me and affected me psychologically. In brief, I felt exposed to everyone who was passing in that delivery area*.”

The pandemic shaped participants’ comfort with the health facility environment such that over half felt concerned about COVID-19 transmission at the health facility. This concern stemmed from a general worry about the danger of this virus and feeling that the risk of exposure was high at the health facility. Those who said they were unconcerned explained that they felt comfortable due to the prevention measures in place and adherence to these measures.

Many respondents reported being screened for fever upon arrival at the health facility, but none were tested for COVID-19. All reported that providers were wearing personal protective equipment and that prevention measures such as masking, social distancing and handwashing were in place. A few respondents remarked that the maternity unit was crowded, and a couple explained that “*after delivery, the midwife put two women in each bed*,” violating social distancing. This made a respondent feel particularly uncomfortable as she “*was afraid of being infected by COVID-19*.”

##### Facility culture

There were no reports of providers making unreasonable demands of participants, such as requesting they clean up their own bodily fluids. Most participants reported no challenges related to payment for services with some explaining “*it is for free.”* There were still some reports of extortion whereby respondents described being ignored by providers, receiving substandard care, or being in conflict with providers, until they could make payment. One participant explains:

> *I was held up due to inability to pay money to some midwives. But, after some time I paid the money and services got better. In some health facilities when you don’t pay some money to the midwives, they never treat very well.*

Several participants reported that the COVID-19 pandemic negatively affected their ability to save and pay for transport or service costs related to delivery. They described facing economic hardship including loss of work, loss of wages, and increasing prices. For example, one participant shared:

> *We didn’t face any constraint about transport because we have a car in our house, but for delivery services, that was a sacrifice for us. […] I lost my job due to COVID-19 and the situation was very hard for me.*

In one case, the intention to uphold COVID-19 prevention measures became a barrier to care. A participant who experienced a precipitous birth and delivered *en route* to the health facility recounted that she and her husband were initially denied entry to the health facility as they did not have masks with them:

> *After arriving in the main gate of health facility, the security officer demanded us to wear masks, but we didn’t have any masks, and suddenly my husband entered the health facility by force and the health staff gave us all necessary support.*

### Satisfaction

The majority of participants said they felt satisfied with the services they received, with only one outright indicating dissatisfaction. Most explained they felt satisfied with their experience “*Because I delivered my baby without any constraint,”* referring to delivering without any major complications.

Several shared that they felt satisfied because of the positive interactions they had with providers, as described by one participant saying, “*I was extremely happy because I had achieved what I was expecting about delivery services. The midwives were entirely committed with their service in order to help me*.” One respondent who had described major incidents of mistreatment still said she felt very satisfied with the services, explaining,

> *That was excellent for me because of the experience of delivering my second baby who is healthy and beautiful. Regarding the second midwife, it is a situation which regularly occurs in our health facilities, so I left it behind.*

## Discussion

Overall, this study demonstrated that COVID-19 negatively affected some aspects of maternal and neonatal quality of care, however, the effects were limited. There were some cases in which pandemic-related adaptations restricted involvement of birth companions, limiting social support and countering cultural practices, and one case in which a family was initially prevented from accessing care. Pre-existing conditions of overcrowding became more worrisome to clients during the pandemic due to the fear of infection transmission. These scenarios suggest a need for the development of cohesive policies and appropriate enforcement of such policies that balance the need to address shocks to the health system with the need to continue delivering high quality person-centered care regardless of the circumstance. At the same time, most of the quality-of-care issues highlighted in this research, including disrespect, appear to be chronic health system struggles, not necessarily associated with, or exacerbated by, the pandemic.

The results are consistent with previous research documenting disrespectful care in Mozambique prior to the pandemic. One study conducted in two Maputo City district hospitals showed widespread disrespect and abuse during childbirth with high levels of non-confidential care, non-consented care, and abandonment.(31) Similarly, this study found widespread unconsented care for both women and newborns and some cases of abandonment. This study adds to the research by identifying that unconsented care is normalized in this context. Another study in southern Mozambique demonstrated that birthing persons who experienced disrespect and abuse in childbirth reported lower levels of satisfaction with their care, which aligns with the findings in the current study whereby participants linked health service satisfaction with respectful care.(32)

The current study results highlight several gaps in the provision and experience of care that should be considered for intervention. First, participant reports of screening at admissions suggest a missed opportunity. Skipping simple questions about danger signs and symptoms during intake can delay diagnosis and treatment of major maternal and newborn complications. This challenge has similarly been documented by other studies and across settings.(33–35) Health planners might explore the use of an admissions checklist to ensure these critical, but easily bypassed steps, are consistently conducted.

Second, the frequent reports of non-evidence-based interventions such as episiotomies and enemas are concerning. Episiotomy can be a useful technique, however, professional associations such as the International Federation of Gynecology and Obstetrics (FIGO) and the American College of Obstetricians and Gynecologists (ACOG) recommend “judicious” and “restrictive” use rather than routine application.(30, 36, 37) The benefits of episiotomy must be carefully weighed against the increased risks of blood loss, depth of posterior perineal injury, anal sphincter damage, increased pain, and long-term urinary incontinence.(38–40) The many reports of episiotomy in this study indicate a need to investigate the prevalence of this practice and to support providers in alternative management practices, as possible, to reduce use of episiotomy.

Similarly, reports of enemas were commonplace in this study even though there is no evidence to support any benefits of routine enema during the first stage of labor.(41) Routine rectal enema is deemed an unnecessary intervention that may cause discomfort to clients, increase the workload of providers and increase the cost to the health system.(41) While enema is not recommended, many participants in this study noted they were favorable towards the procedure, making statements suggesting that defecation during delivery was not socially acceptable. Women should certainly be supported in their health care decisions, however, it is important to ensure they are truly given a choice in their health care. A practice of routine enemas and related social norms can create an environment in which women feel shame about their natural bodily response to defecate during labor leading them to feel disempowered to decline the procedure. Actions may be needed to reduce such stigma to ensure women have a full range of choice in their birth care options that allow for bodily autonomy.

Third, there was a considerable gap in offering pain management. This is unsurprising considering the shortage of anesthesiologists and anesthesia technicians in Mozambique, who are needed to administer common pain management options in labor such as epidural, inhaled analgesia and local anesthetic nerve block.(42) Some participants were offered “pills,” however, oral non-opioid medications offer limited pain relief and there is little evidence to support their effectiveness in reducing labor pain.(43) While some participants expressed appreciation for the pills they received and described it as a contributor to their birth satisfaction, this is more likely related to having their concerns heard rather than a reduction in physical pain. There are of course non-pharmacological interventions that can be used to manage discomfort in labor. There is evidence indicating that massage, relaxation techniques and immersion in water can reduce pain during labor.(43) In addition, research has found that continuous labor support (for example, of a doula or labor companion) is associated with a decreased likelihood in the use of intrapartum analgesia, suggesting that birth companions may assist with pain management.(44) Indeed, the current study documented several examples of nurses and birth companions offering comfort measures to manage labor pains. In a context of health workforce constraints and commodity stockouts, more widespread dissemination of non-pharmacological approaches to pain management could provide women with at least one option for effective pain relief. Offering doula training to traditional birth attendants could fill this important gap while simultaneously allowing for better integration of traditional birth attendants in the health system.

Overall, both positive and negative experiences of care were reported in this research. While there is mounting evidence that disrespect and abuse during childbirth exists and at unacceptable levels, there is a need to also capture *respectful* care in action. Maternity providers face tremendous challenges in their work including lack of decision-making power and autonomy, unsafe work environments, gender inequities, violence, under- and inconsistent pay, lack of mental health support, understaffing and unsustainable work hours, among others.(45) From this, it is clear that efforts to improve respectful maternity care must address these health systems issues. Nevertheless, there is a role for provider behavior change to achieve respectful care. To enable this behavior change, it will be helpful, and potentially more effective, to offer providers positive examples of care that can be modeled and emulated rather than emphasizing only the problematic behaviors.

The many positive care experiences shared in this study offer hope, however, we must underscore the serious mistreatment of women during childbirth that was also recorded. Globally, as there is well-documented gender bias in health care and minimization of women’s experience of pain, it is critical to keep monitoring disrespectful maternity care. Participants in this study experienced threats of cesarean section, sexism towards their newborns, suturing without anesthesia, non-evidence-based care, neglect, extortion, and unconsented care. These represent a violation of human rights and, fundamentally, of human dignity. We urge health system actors in Mozambique to take swift action to uphold the dignity of all women.

There are a few limitations that should be considered when interpreting the results of this study. First, this study does not have a pre-pandemic comparison, making it difficult to disentangle the extent to which the COVID-19 pandemic may have contributed to pre-existing quality of care challenges. Further research documenting the health sector experience during the pandemic in Mozambique may help elucidate any shifts in quality of care that are less visible to clients of the system. Second, the focus of this study was on non-surgical live births. As such, additional quality issues associated with surgery and maternal and newborn mortality have not been captured here. The aim of the current study, however, was to document the common experience of birth during the pandemic, which is mainly represented by those with a non-surgical live birth. Even with this focus, the research readily identified mistreatment during childbirth, indicating a substantial need to improve routine intrapartum care. Third, this study captures self-reported experiences of care which may be affected by social desirability bias. At the same time, recent research comparing labor observations to postpartum client self-report found comparable prevalence estimates for most forms of mistreatment.(46) Further, in taking a person-centered approach to health care, it is important to consider self-report as the reference standard. Finally, provider recruitment of participants may have invoked some selection bias. However, clients were recruited at their first ANC visit when the later labor experience was yet unknown, and the recruiting provider would not necessarily be involved in the client’s birth support.

There are several key implications from the findings. First, as many participants were surprised by the respectful, quality care they received, there is a need to better communicate health system improvements to the community, which may help increase uptake of institutional delivery. Second, documenting client birth satisfaction through patient feedback mechanisms is insufficient since the findings show that in this setting clients largely base satisfaction on birth outcome rather than the full gamut of their experience of care. To gain insight into client experience, more nuanced questions are required about interactions with providers, procedures received and the conditions of the health facility. Third, it will be important to harness positive examples of care to grow a culture of respectful care. Only highlighting disrespectful care gives way to a punitive environment that encourages secrecy about mistreatment. Given respectful care is already being practiced, recognizing and rewarding this behavior may be more effective in fostering organizational norms of respectful care than focusing on negative behavior. Fourth, some forms of disrespect and mistreatment have been normalized in this context by clients. To meaningfully shift these practices will require a combination of provider behavior change and social norms interventions. Finally, there is a need to invest in health systems approaches to create an enabling environment that can support a shift in organizational culture and social norms.

## Data Availability

Qualitative transcripts will be made available upon reasonable request. Transcripts will not be uploaded to a data repository given the sensitive nature of the interviews and the potential to identify participants by triangulation of responses.

## Acknowledgements

We sincerely appreciate all the participants in this study who generously shared their time and stories with us. We thank you for your courage and willingness to recount your experiences. We are grateful to the FHI 360 Alcançar team and the Reproductive, Maternal, Newborn and Child Health Division at FHI 360 for their support in this effort. Special thanks to Geoffrey Ezepue, Fulgencio Estrada, and Donna McCarraher. We greatly appreciate Holly Burke’s critical review of the draft manuscript.

## Author contributions

MML led study conceptualization and methodology design in collaboration with JV and EK. JV led project administration and supervision of data collection as well as data validation. AB contributed to project administration and data validation. CC and MML performed formal analysis. MML wrote the original draft manuscript with review and editing from CC, AB, JV and EK.

